# Digital Health Technology Burden and Frustration Among Patients with Multimorbidity

**DOI:** 10.1101/2025.10.09.25337645

**Authors:** Haoxin Chen, Jiancheng Ye

## Abstract

**Objective:** This study aimed to characterize patterns of digital health technology (DHT) use and examine the relationships between multimorbidity, DHT adoption, and user-reported frustration, with a focus on identifying socioeconomic and phenotypic determinants of digital health burden.

**Methods:** A cross-sectional analysis was conducted using nationally representative public data from HINTS 7 (2024). Adults with at least one chronic condition were included (N=3,753). The outcomes were the number of DHTs used and frustration with digital tools. Multivariable logistic regression models were employed, adjusting for sociodemographic and clinical variables. Phenotypic subgroup analyses were conducted based on multimorbidity, DHT use, and frustration patterns.

**Result:** Among 3,753 participants, 46.9% had multimorbidity. Participants with multimorbidity reported using a greater number of DHTs on average (mean = 3.9 vs. 3.6, p < 0.001) compared to those with a single condition, yet exhibited significantly higher rates of frustration with digital tasks (61.2% vs. 54.1%, p < 0.001). Both higher income and educational groups were associated with lower odds of frustration. Greater DHT use was also independently associated with reduced frustration (OR = 0.78, 95% CI: 0.71-0.87, p < 0.01). Phenotypic subgroup analysis further identified individuals with multimorbidity and low DHT use as the most vulnerable profile, characterized by older age, lower socioeconomic status, and the highest frustration prevalence (60.3%).

**Conclusion:** While individuals with multimorbidity use more DHTs, they experience greater frustration, particularly those with lower socioeconomic status. However, higher engagement with DHTs is associated with lower frustration, suggesting that technology proficiency may mitigate burden. Targeted interventions addressing digital literacy and user-centered design are needed to experience among patients with complex chronic conditions.

## INTRODUCTION

Digital health technologies (DHTs), including patient portals, telehealth platforms, mobile health applications, and wearable devices, are becoming deeply embedded in the infrastructure of chronic disease management. While these tools have the potential to expand access, support self-management, and enable remote monitoring, their real-world impact on patient workload, experience, and outcomes remains inconsistent and complex.[1] This complexity is particularly evident among the growing population living with multimorbidity - defined as having two or more chronic conditions. These individuals navigate inherently complex care regimens, experience fragmented care delivery, and shoulder substantial self-management responsibilities.[2] The layering of multiple, often condition-specific DHTs onto these existing demands creates a critical juncture that may either streamline care coordination and empower patients, or unintentionally amplify their treatment burden.[3] Understanding the determinants of these divergent pathways, where DHTs facilitate versus hinder care, is therefore important to achieving patient-centered implementation.

Emerging evidence demonstrates that DHTs can improve selected clinical outcomes and care processes in specific chronic conditions.[4] However, these benefits are not universal. Effects are heterogeneous and contingent on factors such as usability, integration into clinical workflows, and surrounding support models.[5, 6] Reviews of digital interventions and implementation efforts in chronic disease management highlight benefits for patient engagement and provider coordination, yet also underscore barriers such as limited digital literacy, poor workflow fit, and fragmented digital ecosystems that shift administrative work from clinics to patients and families.[7] Despite this growing recognition of digital barriers, a substantial gap persists. Few studies have systematically quantified patient-reported frustration with DHTs or identified specific vulnerability phenotypes using large, nationally representative datasets. This leaves an insufficient understanding of how these digital barriers manifest and are distributed across the population, particularly among patients with complex chronic care needs.[8] The Burden of Treatment (BoT) framework offers a valuable lens through which to examine this issue.[9] It conceptualizes health-related work as a function of the workload imposed by healthcare (e.g., learning new technologies, monitoring symptoms, scheduling virtual appointments) and the capacity individuals have to manage it (e.g., skills, resources, social support).[7] For individuals with multimorbidity, who often juggle multiple portals, devices, and applications, the workload imposed by DHTs can be substantial, especially when technologies are numerous, functionally overlapping, poorly integrated, or misaligned with their needs and capacities.[10] This raises a pressing concern that the rapid expansion of DHTs, if not designed and implemented with equity as a core principle, risks widening existing health disparities by failing to meet the needs of the most vulnerable groups. A recent scoping review focused on multimorbidity have called for more standardized approaches to characterize the types and intensity of DHT use, document patients’ negative experiences, and evaluate the overall balance of benefits versus burdens.[8]

To address these complex questions, this study adopts a methodological approach aligned with contemporary public health research that leverages nationally representative surveys to characterize disease prevalence and identify population phenotypes. This paradigm is widely employed in epidemiological studies, such as those utilizing the National Health and Nutrition Examination Survey (NHANES) to define the prevalence and subclassifications of metabolic dysfunction-associated steatotic liver disease under new consensus nomenclature,[11] and to investigate the relationship between serum biomarkers and obesity phenotypes using multivariable logistic regression and restricted cubic splines.[12] Similarly, the use of large-scale longitudinal cohorts such as the China Health and Retirement Longitudinal Study (CHARLS) to examine the association between sarcopenia and frailty through multivariable regression and subgroup analysis demonstrates the robustness of such methods for identifying at-risk subgroups in aging populations.[13]

Building upon these foundations, this study aims to comprehensively assess DHT usage and burden among adults with multimorbidity using a nationally representative dataset. We first quantify the number and types of DHTs used by this population, then model the association between multimorbidity status and DHT use, while simultaneously examining the association with patient-reported frustration. Furthermore, we conduct phenotypic analyses through stratification to identify vulnerable subgroups. This research seeks to generate evidence that can inform pragmatic, equity-minded strategies to optimize digital care for people with multiple chronic conditions, maximizing the benefits of technology while minimizing associated workload.

## METHODS

### Study design

We conducted a cross-sectional analysis of data from the Health Information National Trends Survey (HINTS 7)[14] from the National Cancer Institute. HINTS surveys are administered biennially among noninstitutionalized US civilians aged ≥18 years and contain nationally representative data on awareness and use of health-related information. The HINTS 7 sample comprised 7,278 respondents, with surveys conducted from March 25, 2024, to September 16, 2024. We excluded observations with missing, erroneous, or inapplicable values, resulting in a final analytical sample of 3,753 participants.

### Measures

Multimorbidity was operationalized based on a count of self-reported chronic conditions, including diabetes, hypertension, heart disease, and lung disease. Participants reporting zero chronic conditions were excluded from subsequent analyses. DHT use was assessed through survey items covering several domains: patient portal functions (e.g., information lookup, secure messaging, accessing test results, and appointment scheduling), health and wellness applications, organizer or portal apps, wearable devices, and sharing of device-generated data with healthcare providers. Responses were summed to create a continuous variable ranging from 0 to 8, representing the total number of distinct technology types used. Frustration with DHTs was measured by recoding agreement with the statement that digital tasks are frustrating into a binary variable, where 1 indicated “somewhat agree” or “strongly agree” and 0 indicated “somewhat disagree” or “strongly disagree.”.

Key sociodemographic covariates were included in the analysis: age, sex, race/ethnicity, education, household income, and health insurance status. To enhance model stability and interpretability, education and income were recategorized into three ordinal levels. Education was grouped as: (1) high school or less, (2) some college or associate degree, and (3) bachelor’s degree or higher. Income was categorized as: (1) less than $20,000, (2) $20,000 to $74,999, and (3) $75,000 or more. All categorical variables were dummy coded with explicit reference categories specified in the regression models.

### Descriptive analysis

Descriptive analyses were conducted to summarize the prevalence of multimorbidity, the distribution of DHT use, and participant demographics. Continuous variables are presented as mean ± standard deviation (SD) and compared using independent samples t-tests. Categorical variables are reported as frequencies and percentages [n (%)] and compared using chi-square tests. Statistical significance was defined as p < 0.05 for all analyses.

### Regression models

Two multivariable logistic regression models were employed. Model 1 assessed the association between DHT use, frustration with DHTs, and multimorbidity status, adjusted for sociodemographic factors. Specifically, a logistic regression model was fitted with multimorbidity as the outcome, regressed on age, income, education, health insurance status, sex, race/ethnicity, the number of DHTs used, and frustration with DHTs. This model estimated adjusted odds ratios (aORs) with 95% confidence intervals (CIs) for the factors associated with having multimorbidity.

Multimorbidity ∼ Age + Income + Education + Insurance + Sex + Race/Ethnicity + Number of DHTs + Frustration (Model 1)

Model 2 evaluated the association between multimorbidity status, DHT use, and frustration with digital tasks, controlling for the same set of covariates. Using logistic regression, frustration (binary: 0 = not frustrated, 1 = frustrated) was the outcome, with multimorbidity status, the number of DHTs used, and all sociodemographic variables included as independent variables. The aORs with corresponding 95% CIs were computed to quantify these relationships.

Frustration ∼ Multimorbidity + Number of DHTs + Age + Income + Education + Insurance + Sex + Race/Ethnicity (Model 2)

For both models, multicollinearity was assessed using variance inflation factors (VIF), and model fit was evaluated using the Hosmer-Lemeshow goodness-of-fit test.

### Subgroup analysis

To further explore the interplay between multimorbidity, DHT use, and user frustration, we conducted subgroup analyses based on two distinct 2×2 grouping strategies to identify clinically relevant phenotypes.

First grouping strategy: Participants were classified by multimorbidity status (single vs. multiple conditions) and level of DHT adoption. We used a cutoff of 6 technologies to define higher use (≥ 6 DHTs) versus lower use (<6 DHTs). This threshold was determined based on the distribution of technology adoption in the study population, corresponding to the 75^th^ percentile of DHT use. The outcome of interest for this classification was the presence of frustration with DHTs. Differences in frustration levels and other key characteristics across the resulting four groups were assessed using one-way ANOVA for continuous variables (with post-hoc Tukey tests where appropriate) or Kruskal-Wallis tests for non-normally distributed data, and chi-square tests for categorical variables.

Second grouping strategy: Participants were categorized by multimorbidity status (single vs. multiple conditions) and the presence or absence of frustration (binary: not frustrated vs. frustrated). For this stratification, the outcome was the level of DHT use. Group differences in the number of technologies utilized, along with other sociodemographic variables, were evaluated using one-way ANOVA or Kruskal-Wallis tests as appropriate.

For both grouping approaches, we profiled the resulting subgroups by examining the distribution of sociodemographic characteristics. Additionally, to visually represent patterns of specific DHT usage frequencies across the four groups within each classification, radar plots were generated using normalized scores, providing a comparative overview of technology engagement behaviors across multiple dimensions of digital health literacy and operational competence.

### Study Approval

We solely used publicly available HINTS data for this study.

## RESULTS

### Descriptive Analysis

Descriptive statistics for the study participants, stratified by multimorbidity status, are presented in **Table 1**. The analytical sample comprised 1,990 individuals with a single chronic condition and 1,763 individuals with multimorbidity (two or more conditions), representing 53.1% and 46.9% of the sample, respectively.

**Table 1.** Characteristics of study participants stratified by multimorbidity status.

| Variables | Total<br>N = 3753 | One<br>condition<br>n = 1990 | Two or more<br>conditions<br>n = 1763 | p-value |
| --- | --- | --- | --- | --- |
| Age, years, Mean (SD) | 60.0 ± 20.2 | 57.8 ± 20.6 | 62.5 ± 19.6 | **<0.001** |
| Number of conditions, Mean (SD) | 1.6 ± 0.8 | 1.0 ± 0.0 | 2.3 ± 0.6 | **<0.001** |
| Types of technology used, Mean (SD) | 3.7 ± 2.2 | 3.9 ± 2.1 | 3.6 ± 2.3 | **<0.001** |
| Income ranges, N (%) |  |  |  | **<0.001** |
| \$0 - \$19,999 | 720 (21.0) | 315 (17.2) | 405 (25.3) | |
| \$20,000 - \$74,999 | 1509 (44.0) | 767 (42.0) | 742 (46.3) | |
| $\geq \$75,000$ | 1199 (35.0) | 745 (40.8) | 454 (28.4) | |
| Education, N (%) |  |  |  | **<0.001** |
| High school or lower | 951 (26.2) | 445 (23.1) | 506 (29.7) |  |
| High school graduate/some college | 1180 (32.5) | 592 (30.7) | 588 (34.6) |  |
| Bachelor or higher | 1498 (41.3) | 890 (46.2) | 608 (35.7) |  |
| Race, N (%) |  |  |  | **<0.001** |
| Hispanic | 606 (17.6) | 336 (18.3) | 270 (16.8) |  |
| Asian | 142 (4.1) | 84 (4.6) | 58 (3.6) |  |
| Black | 656 (19.1) | 307 (16.7) | 349 (21.7) |  |
| White | 1920 (55.8) | 1058 (57.5) | 862 (53.7) |  |
| Other | 120 (3.5) | 54 (2.9) | 66 (4.1) |  |
| Sex, N (%) |  |  |  | 0.056 |
| Female | 2119 (58.3) | 1153 (59.8) | 966 (56.6) |  |
| Male | 1517 (41.7) | 776 (40.2) | 741 (43.4) |  |
| Health insurance, N (%) | 3494 (93.1) | 1834 (92.2) | 1660 (94.2) | **0.038** |
| Diabetes, N (%) | 1521 (40.5) | 284 (14.3) | 1237 (70.2) | **<0.001** |
| High blood pressure, N (%) | 2953 (78.7) | 1322 (66.4) | 1631 (92.5) | **<0.001** |
| Heart condition, N (%) | 720 (19.2) | 86 (4.3) | 634 (36.0) | **<0.001** |
| Lung disease, N (%) | 914 (24.4) | 298 (15.0) | 616 (35.0) | **<0.001** |

Significant differences were observed between the two groups across sociodemographic, healthcare access, and technology-related variables. Participants with multimorbidity were significantly older than those with a single condition (mean [SD] age: 62.5 ± 19.6 years vs. 57.8 ± 20.6 years, p < 0.001). Racial composition differed significantly between groups (p < 0.001); the proportion of Black individuals was higher in the multimorbidity group compared to the single condition group (21.7% vs. 16.7%), while the proportion of White individuals was correspondingly lower (53.7% vs. 57.5%). Income distribution differed significantly, with a higher prevalence of high household income (≥ $75,000) observed in the single condition group compared to the multimorbidity group (40.8% vs. 28.4%, p < 0.001). Education level also varied markedly, as participants with a single condition exhibited a lower proportion of individuals with high school education or less (23.1% vs. 29.7%, p < 0.001) and a correspondingly higher proportion with bachelor’s degrees or higher (46.2% vs. 35.7%).

Regarding DHT use, individuals with a single chronic condition reported using fewer types of technologies compared to those with multimorbidity (mean = 3.6 ± 2.1 vs. 3.9 ± 2.3, p < 0.001). Significant differences were also observed in health insurance coverage, with a slightly higher proportion of insured individuals in the multimorbidity group (94.2% vs. 92.2%, p = 0.038). The prevalence of specific chronic conditions differed substantially between groups, with the multimorbidity group showing higher rates of diabetes (70.2% vs. 14.3%, p < 0.001), high blood pressure (92.5% vs. 66.4%, p < 0.001), heart conditions (36.0% vs. 4.3%, p < 0.001), and lung disease (35.0% vs. 15.0%, p < 0.001).

**Table 2** presents detailed DHT usage patterns stratified by multimorbidity status. In terms of specific DHT activities, the single condition group demonstrated higher rates of using the internet for health information (83.7% vs. 79.6%, p = 0.002), making appointments online (65.7% vs. 59.1%, p < 0.001), using wellness apps (52.9% vs. 48.8%, p = 0.016), and using wearable devices to track health (34.8% vs. 29.7%, p < 0.001). Conversely, individuals with multimorbidity were more likely to share health device information with healthcare providers (32.2% vs. 22.2%, p < 0.001). The single condition group reported higher daily social media usage (55.9% vs. 48.8%, p < 0.001) and lower rates of never using social media (18.8% vs. 24.8%). Similarly, they were more likely to watch health-related videos on social media (p < 0.001). Regarding technology-related frustration, the multimorbidity group had a higher proportion strongly agreeing with feeling frustrated (22.5% vs. 16.4%, p < 0.001) and a lower percentage strongly disagreeing (16.0% vs. 22.0%).

**Table 2.** Digital health technology usage patterns and social media engagement stratified by multimorbidity status.

| Variables | Total<br>N = 3753 | One<br>condition<br>n = 1990 | Two or more<br>conditions<br>n = 1763 | p-value |
| --- | --- | --- | --- | --- |
| Used internet for health information, N (%) | 2846 (81.8) | 1574<br>(83.7) | 1272 (79.6) | **0.002** |
| Used internet to send message, N (%) | 2275 (65.5) | 1249<br>(66.6) | 1026 (64.3) | 0.175 |
| Used internet to view results, N (%) | 2580 (74.4) | 1404<br>(74.9) | 1176 (73.9) | 0.522 |
| Used internet to make appointment, N (%) | 2175 (62.4) | 1233<br>(65.7) | 942 (59.1) | **<0.001** |
| Used wellness app, N (%) | 1808 (50.9) | 1006<br>(52.9) | 802 (48.8) | **0.016** |
| Used portal organizer app, N (%) | 195 (5.3) | 96 (4.9) | 99 (5.7) | 0.299 |
| Used wearable device to track health, N (%) | 1200 (32.3) | 685 (34.8) | 515 (29.7) | **<0.001** |
| Shared health device information, N (%) | 984 (27.2) | 433 (22.2) | 551 (32.2) | **<0.001** |
| Visited social media, N (%) |  |  |  | **<0.001** |
| Almost every day | 1946 (52.5) | 1098 (55.9) | 848 (48.8) |  |
| At least once a week | 432 (11.7) | 225 (11.5) | 207 (11.9) |  |
| A few times a month | 286 (7.7) | 156 (7.9) | 130 (7.5) |  |
| Less than once a month | 239 (6.5) | 117 (6.0) | 122 (7.0) |  |
| Never | 801 (21.6) | 369 (18.8) | 432 (24.8) |  |
| Shared personal health information on social media, N (%) |  |  |  | 0.789 |
| Almost every day | 58 (1.6) | 28 (1.4) | 30 (1.7) |  |
| At least once a week | 78 (2.1) | 37 (1.9) | 41 (2.4) |  |
| A few times a month | 112 (3.0) | 58 (3.0) | 54 (3.1) |  |
| Less than once a month | 342 (9.3) | 183 (9.4) | 159 (9.2) |  |
| Never | 3096 (84.0) | 1651 (84.4) | 1445 (83.6) |  |
| Shared general health information on social media, N (%) |  |  |  | 0.098 |
| Almost every day | 55 (1.5) | 25 (1.3) | 30 (1.7) |  |
| At least once a week | 103 (2.8) | 45 (2.3) | 58 (3.4) |  |
| A few times a month | 221 (6.0) | 116 (5.9) | 105 (6.1) |  |
| Less than once a month | 642 (17.4) | 362 (18.5) | 280 (16.2) |  |
| Never | 2664 (72.3) | 1408 (72.0) | 1256 (72.6) |  |
| Interacted with similar people on social media, N (%) |  |  |  | 0.107 |
| Almost every day | 57 (1.5) | 29 (1.5) | 28 (1.6) |  |
| At least once a week | 125 (3.4) | 61 (3.1) | 64 (3.7) |  |
| A few times a month | 204 (5.5) | 96 (4.9) | 108 (6.2) |  |
| Less than once a month | 452 (12.2) | 260 (13.3) | 192 (11.0) |  |
| Never | 2858 (77.3) | 1511 (77.2) | 1347 (77.5) |  |
| Watched health video on social media, N (%) |  |  |  | **<0.001** |
| Almost every day | 144 (3.9) | 75 (3.8) | 69 (4.0) |  |
| At least once a week | 334 (9.0) | 183 (9.3) | 151 (8.7) |  |
| A few times a month | 642 (17.3) | 357 (18.2) | 285 (16.4) |  |
| Less than once a month | 1057 (28.6) | 608 (31.0) | 449 (25.8) |  |
| Never | 1524 (41.2) | 737 (37.6) | 787 (45.2) |  |
| Felt frustrating using new technology, N (%) |  |  |  | **<0.001** |
| Strongly agree | 713 (19.3) | 323 (16.4) | 390 (22.5) |  |
| Somewhat agree | 1413 (38.2) | 742 (37.7) | 671 (38.7) |  |
| Somewhat disagree | 867 (23.4) | 469 (23.8) | 398 (22.9) |  |
| Strongly disagree | 710 (19.2) | 433 (22.0) | 277 (16.0) |  |

However, no significant differences were detected in several other DHT-related activities between the two groups. These included using the internet to send secure messages to providers, viewing test results online, using portal organizer apps, sharing personal and general health information on social media, and interacting with individuals having similar health conditions on social media (p > 0.05). Additionally, sex showed no statistically significant differences between groups (p = 0.056).

### Regression Model Results

The multivariable logistic regression analysis (Model 1) examining factors associated with multimorbidity status are presented in **Figure 1**. After adjusting for all covariates, age and income emerged as statistically significant predictors of multimorbidity. Specifically, each additional year of age was associated with significantly increased odds of having multiple chronic conditions (OR: 1.02, 95% CI: 1.01-1.03, p < 0.001). Participants in the highest income category (≥ $75,000) demonstrated substantially lower odds of multimorbidity compared to those in the lowest income group (< $20,000) (OR: 0.34, 95% CI: 0.21-0.54, p < 0.001). The middle-income group ($20,000-$74,999) also showed lower odds, though to a lesser extent (OR: 0.67, 95% CI: 0.48-0.93, p < 0.05). In contrast, health insurance status, sex, race/ethnicity, and education were not significantly associated with multimorbidity in the fully adjusted model (all p > 0.05). Notably, neither the number of DHTs used nor frustration with DHTs was independently associated with multimorbidity status after adjustment for sociodemographic factors. These findings indicate that older age and lower SES, particularly income level, are the primary demographic risk factors for multimorbidity in this population.

**Figure 1.**
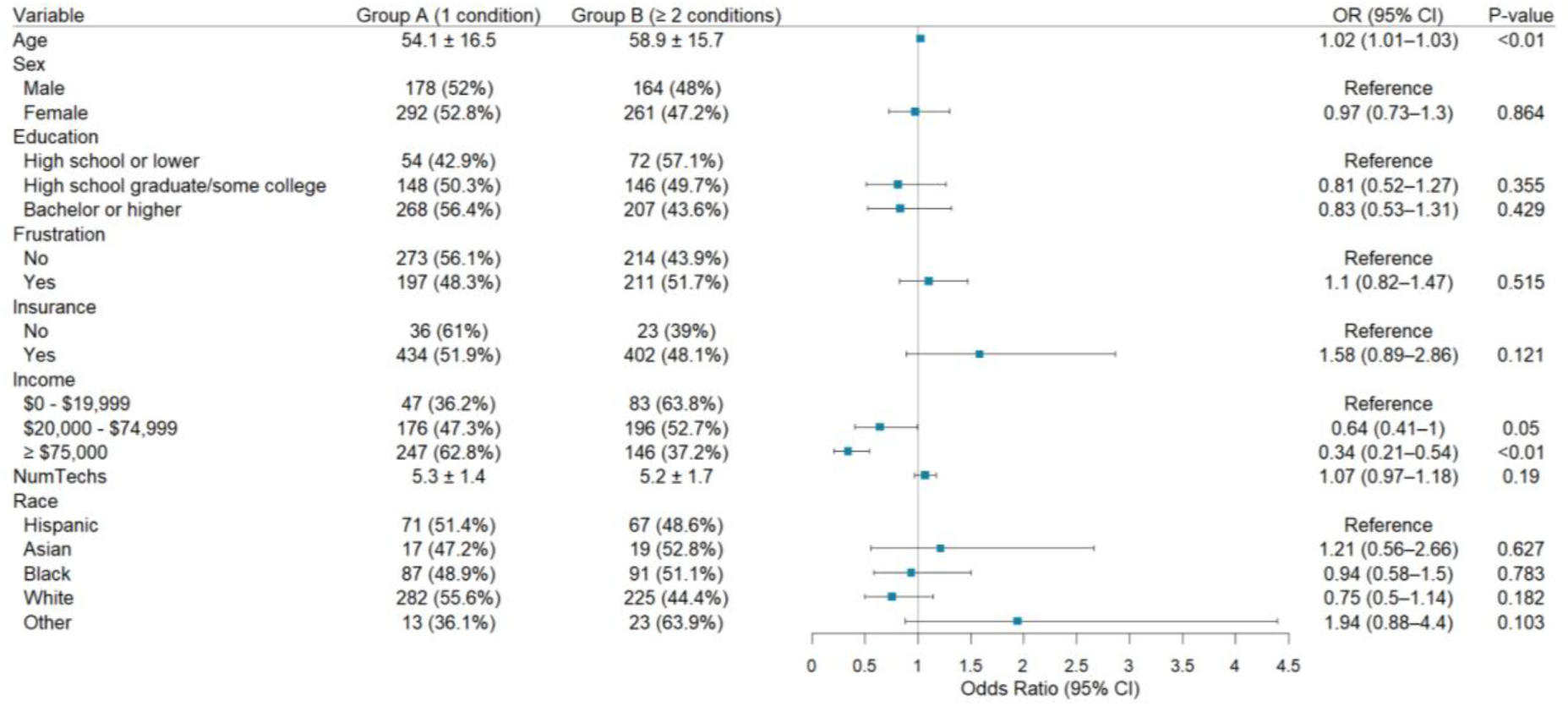
Multivariable logistic regression on predictors of multimorbidity

The results of the multivariable logistic regression examining predictors of frustration with DHTs are shown in **Figure 2**. After adjustment for sociodemographic, clinical, and technology-related factors, several variables demonstrated significant associations with frustration. Older age was associated with slightly increased odds of frustration (OR: 1.02, 95% CI: 1.01-1.03, p < 0.01), with each additional year of age increasing the odds of reporting frustration by approximately 2%. Female was associated with 46% higher odds of frustration relative to male (OR: 1.46, 95% CI: 1.09-1.97, p = 0.013). Participants in the highest income group (≥ $75,000) reported significantly less frustration compared to those in the lowest income category (< $20,000) (OR: 0.50, 95% CI: 0.31-0.80, p < 0.01), representing a 50% reduction in the odds. The middle-income group showed an intermediate effect (OR: 0.72, 95% CI: 0.51-1.01, p = 0.057), approaching but not reaching statistical significance. Similarly, higher education was protective, with bachelor’s degree holders showing 40% lower odds of frustration compared to those with high school education or less (OR: 0.60, 95% CI: 0.42-0.86, p < 0.01). The number of DHTs used was significantly associated with reduced frustration, with each additional technology type associated with 22% lower odds of reporting frustration (OR: 0.78, 95% CI: 0.71-0.87, p < 0.01). The presence of multimorbidity itself was not significantly associated with frustration levels after controlling for other factors (OR: 1.15, 95% CI: 0.89-1.48, p > 0.05). Additionally, no significant associations were observed for race/ethnicity or health insurance status (all p > 0.05).

**Figure 2.**
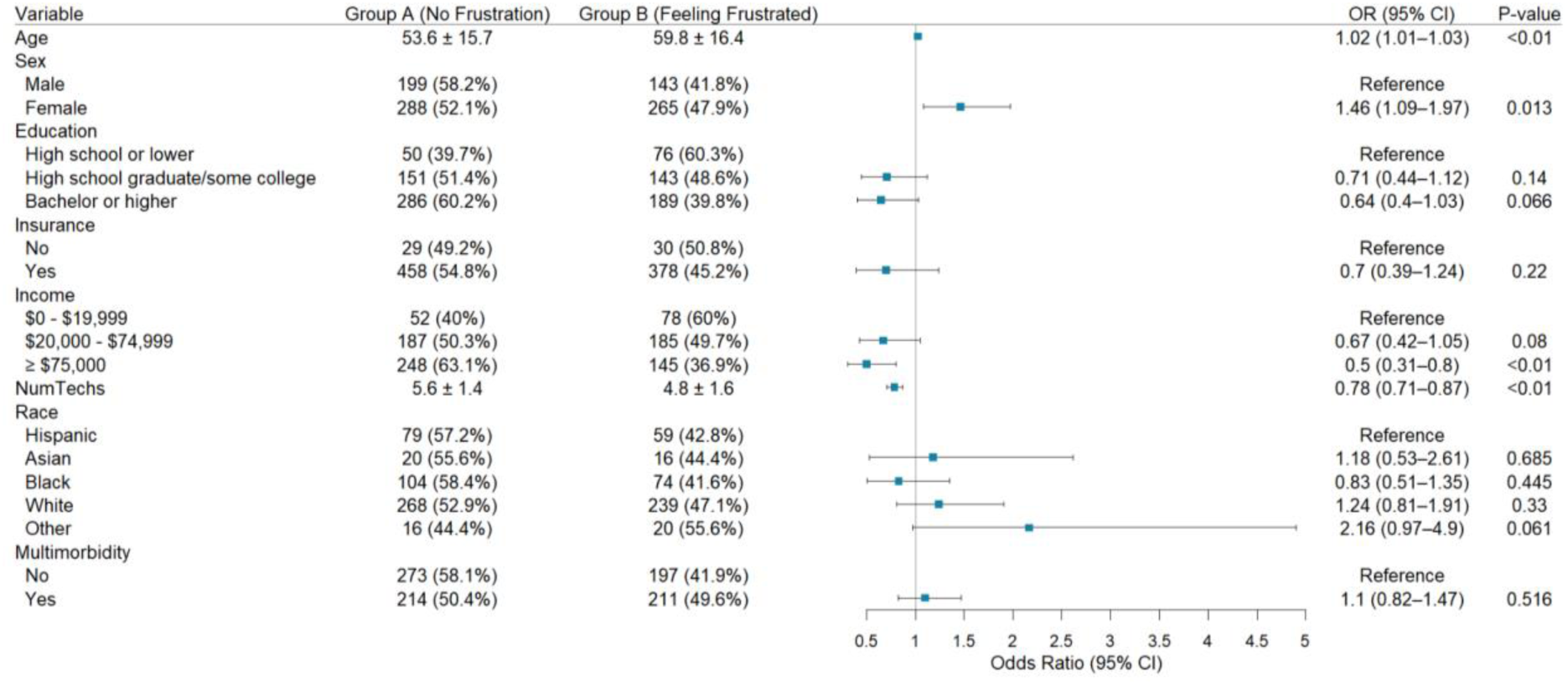
Multivariable logistic regression on predictors of digital health technology frustration

### Subgroup Analysis

#### Subgroup analysis by multimorbidity and DHTs usage

To further elucidate the relationship between multimorbidity, DHT usage intensity, and user frustration, participants were classified into four phenotypic groups based on multimorbidity status (single condition vs. multiple conditions) and level of DHT adoption (<6 technologies vs. ≥6 technologies). A threshold of six technologies was selected to define high DHT use based on the 75^th^ percentile of the distribution, ensuring clinical relevance and adequate sample sizes across groups. Frustration prevalence across these groups was visualized using a 2×2 heatmap (**Figure 3**), and detailed group characteristics are presented in **Table 3**.

**Figure 3.**
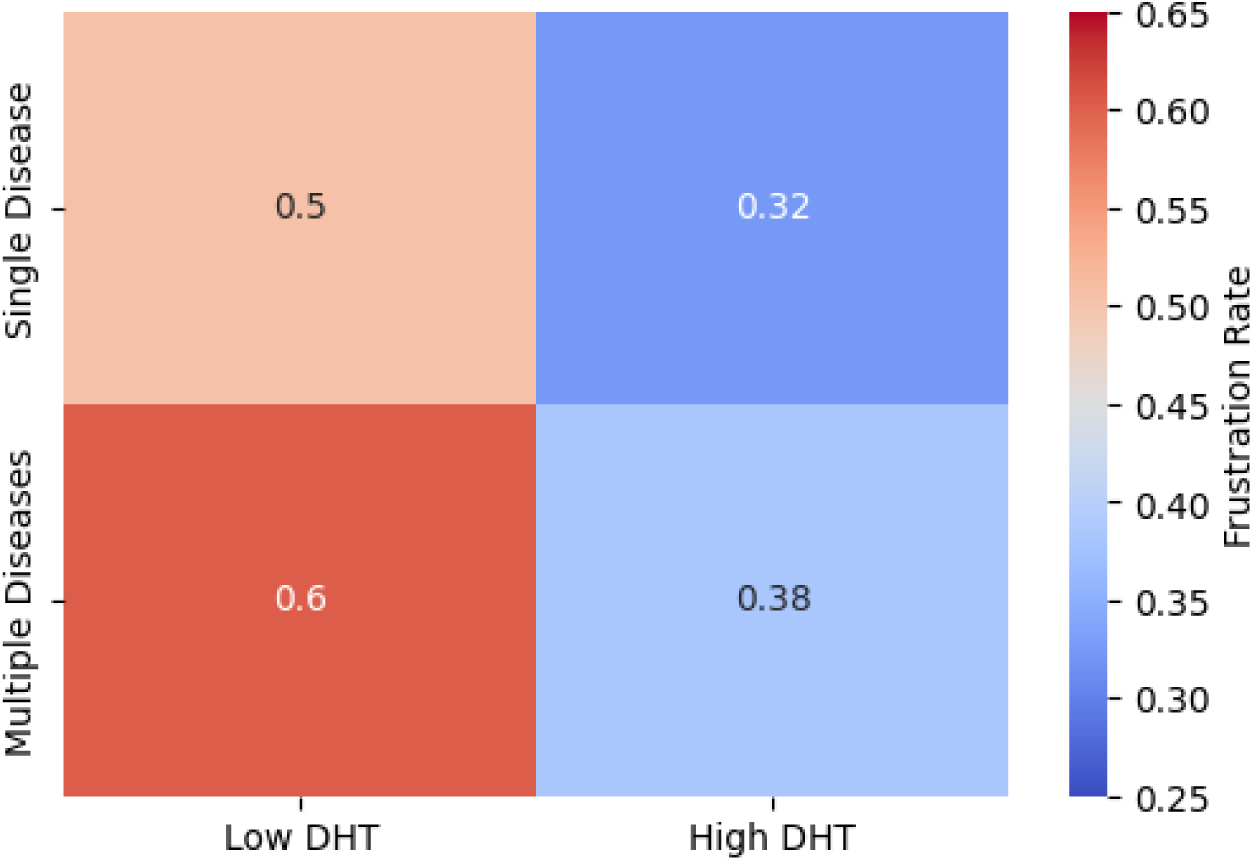
Heatmap of digital health technology frustration prevalence by multimorbidity status and technology usage intensity

**Table 3.** Characteristics of phenotypic subgroups defined by multimorbidity status and digital health technology usage intensity.

| Variable | Group 1<br>n=248 | Group 2<br>n=222 | Group 3<br>n=219 | Group 4<br>n=206 | p-value |
| --- | --- | --- | --- | --- | --- |
| **Age** | 56.7 ± 16.0 | 51.1 ± 16.6 | 61.6 ± 16.3 | 56.1 ± 14.5 | < 0.001 |
| **Frustrated with DHT** |  |  |  |  | < 0.001 |
| No | 123 (49.6%) | 150 (67.6%) | 87 (39.7%) | 127 (61.7%) |  |
| Yes | 125 (50.4%) | 72 (32.4%) | 132 (60.3%) | 79 (38.3%) |  |
| **Income ranges** |  |  |  |  | < 0.001 |
| \$0 - \$19,999 | 27 (10.9%) | 20 (9.0%) | 57 (26.0%) | 26 (12.6%) | |
| \$20,000 - \$74,999 | 110 (44.4%) | 66 (29.7%) | 105 (47.9%) | 91 (44.2%) | |
| ≥ \$75,000 | 111 (44.8%) | 136 (61.3%) | 57 (26.0%) | 89 (43.2%) | |
| **Education** |  |  |  |  | < 0.001 |
| High school or lower | 36 (14.5%) | 18 (8.1%) | 54 (24.7%) | 18 (8.7%) |  |
| High school graduate/some college | 83 (33.5%) | 65 (29.3%) | 85 (38.8%) | 61 (29.6%) |  |
| Bachelor or higher | 129 (52.0%) | 139 (62.6%) | 80 (36.5%) | 127 (61.7%) |  |

The four resulting phenotype groups were:

Group 1: Single condition, low DHT use (n=248)

Group 2: Single condition, high DHT use (n=222)

Group 3: Multiple conditions, low DHT use (=219)

Group 4: Multiple conditions, high DHT use (=206)

Significant differences were observed across the four phenotypes in age, frustration prevalence, income distribution, and education (all p < 0.001). Group 2 was the youngest (mean age 51.1 ± 16.6 years) and exhibited the lowest frustration rate (32.4%). Conversely, Group 3 was the oldest (mean age 61.6 ± 16.3 years) and reported the highest frustration rate (60.3%). Groups 1 and Group 4 demonstrated intermediate and comparable age profiles (56.7 ± 16.0 and 56.1 ± 14.5 years, respectively), though frustration was notably higher in Group 1 (50.4%) than in Group 4 (38.3%). Socioeconomic patterns further distinguish between the groups. Group 2 had the highest proportion of participants with a high income (≥ $75,000: 61.3%) and bachelor’s degree or higher (62.6%). In contrast, Group 3 had the highest concentration of individuals in the lowest income bracket (< $20,000: 26.0%) and the highest proportion with high school education or less (24.7%). Groups 1 and 4 showed intermediate socioeconomic profiles; no significant differences were observed across groups in race/ethnicity, sex, or health insurance status (all p > 0.05). Individuals with multimorbidity but high DHT engagement (Group 4) reported frustration levels comparable to the younger with single conditions and high DHT use (Group 2), suggesting that technology proficiency may mitigate frustration associated with disease complexity. Conversely, those with multiple conditions and low technology use (Group 3) represent a high-risk phenotype characterized by older age, lower socioeconomic status (SES), and the highest digital frustration.

#### Subgroup analysis by multimorbidity and frustration status

Participants were also stratified into four groups based on multimorbidity status (single vs. multiple conditions) and frustration with DHTs (not frustrated vs. frustrated) to examine how the interaction between diseases and technology-related frustration influences the adoption intensity of DHTs. The outcome of interest was high DHT use. Group differences in DHT adoption rates were visualized using the 2×2 heatmap (**Figure 4**), and detailed characteristics are summarized in **Table 4**.

**Figure 4.**
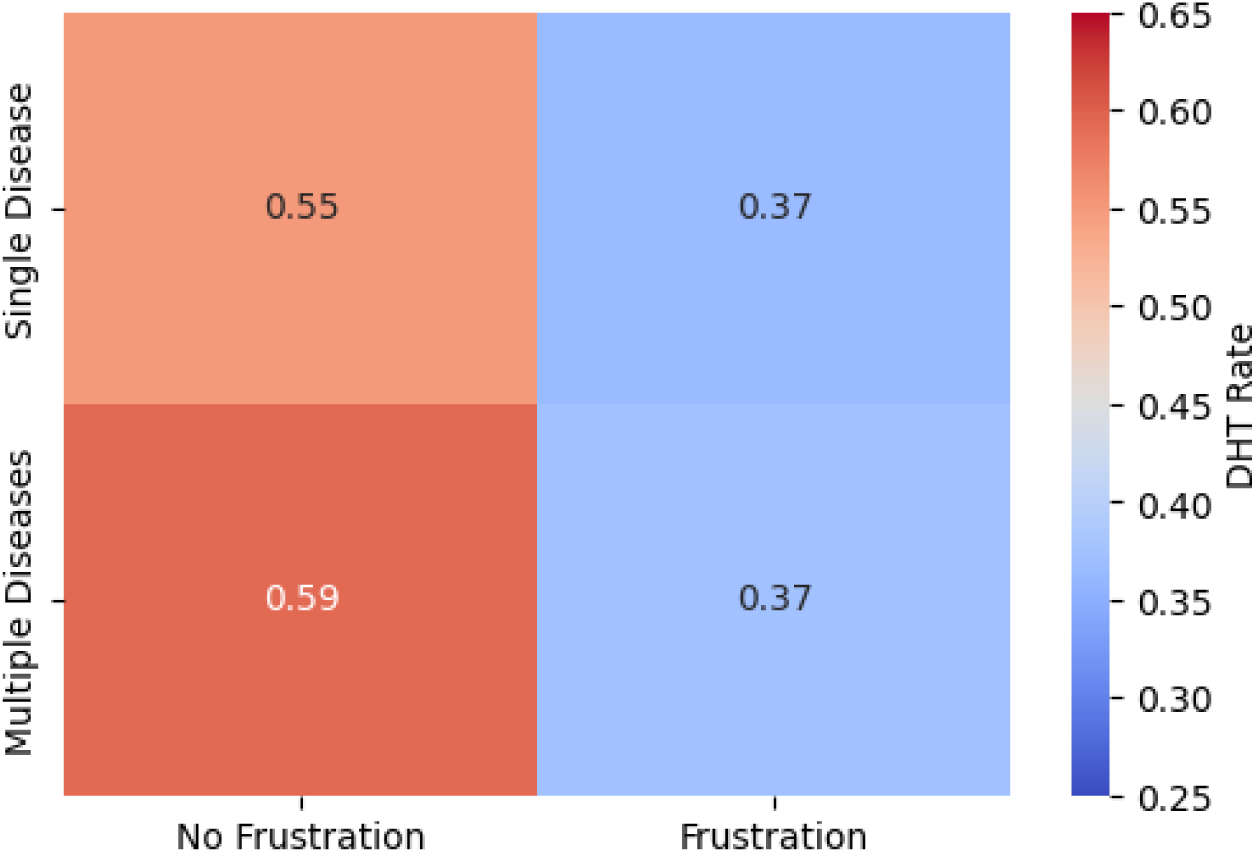
Heatmap of high digital health technology adoption by multimorbidity status and frustration status

**Table 4.** Characteristics of phenotypic subgroups defined by multimorbidity status and digital health technology frustration.

| Variable | Group A<br>n=273 | Group B<br>n=214 | Group C<br>n=211 | Group D<br>n=197 | p-value |
| --- | --- | --- | --- | --- | --- |
| **Age** | 51.8 ± 16.1 | 57.3 ± 16.5 | 55.9 ± 14.8 | 62.1 ± 16.0 | < 0.001 |
| **Frustrated with DHT** |  |  |  |  | < 0.001 |
| No | 123<br>(45.1%) | 125<br>(63.5%) | 87 (40.7%) | 132<br>(62.6%) |  |
| Yes | 150<br>(54.9%) | 72 (36.5%) | 127<br>(59.3%) | 79 (37.4%) |  |
| **Income ranges** |  |  |  |  | < 0.001 |
| \$0 - \$19,999 | 16 (5.9%) | 31 (15.7%) | 36 (16.8%) | 47 (22.3%) | |
| \$20,000 - \$74,999 | 98 (35.9%) | 78 (39.6%) | 89 (41.6%) | 107<br>(50.7%) | |
| ≥ \$75,000 | 159<br>(58.2%) | 88 (44.7%) | 89 (41.6%) | 57 (27.0%) | |
| **Education** |  |  |  |  | 0.0002 |
| High school or lower | 20 (7.3%) | 34 (17.3%) | 30 (14.0%) | 42 (19.9%) |  |
| High school graduate/some college | 85 (31.1%) | 63 (32.0%) | 66 (30.8%) | 80 (37.9%) |  |
| Bachelor or higher | 168<br>(61.5%) | 100<br>(50.8%) | 118<br>(55.1%) | 89 (42.2%) |  |

The four resulting phenotype groups were: Group A: Single condition, not frustrated (n=273) Group B: Single condition, frustrated (n=214) Group C: Multiple conditions, not frustrated (n=211) Group D: Multiple conditions, frustrated (n=197)

Significant differences were observed across the four groups in age, prevalence of high DHT use, income distribution, and education (p < 0.01 for all comparisons). Group A was the youngest (mean age 51.8 ± 16.1 years) and exhibited a high rate of intensive DHT use (55.0% using six or more technologies). Group C, though slightly older (mean age 55.9 ± 14.8 years), demonstrated an even higher level of technology adoption (59.3%), representing the highest DHT engagement rate among all groups. In stark contrast, Group B and Group D showed similarly low rates of high DHT use (36.5% and 37.4%, respectively) despite Group D was substantially older (mean age 62.1 ± 16.0 years) and having greater clinical complexity. Group A had the highest proportion of individuals with high income (58.2%) and bachelor’s degree or higher (61.5%). Group D showed the opposite pattern, with the highest concentration of low-income (22.3% earning < $20,000) and low-education (19.9% with high school or less) individuals, while Groups B and C occupied intermediate positions.

### Digital literacy and operational competence profiles

To further characterize digital health literacy and operational competence across phenotypic subgroups, two radar charts were generated to visualize self-reported confidence and performance across three key dimensions: (1) the ability to use health-related apps without requesting assistance, (2) possessing the skills necessary to find reliable health information online, and (3) experiencing no technical problems during telehealth visits. Results are displayed in **Figure 5a** (grouped by multimorbidity and frustration status) and **Figure 5b** (grouped by multimorbidity and DHT usage level).

**Figure 5a.**
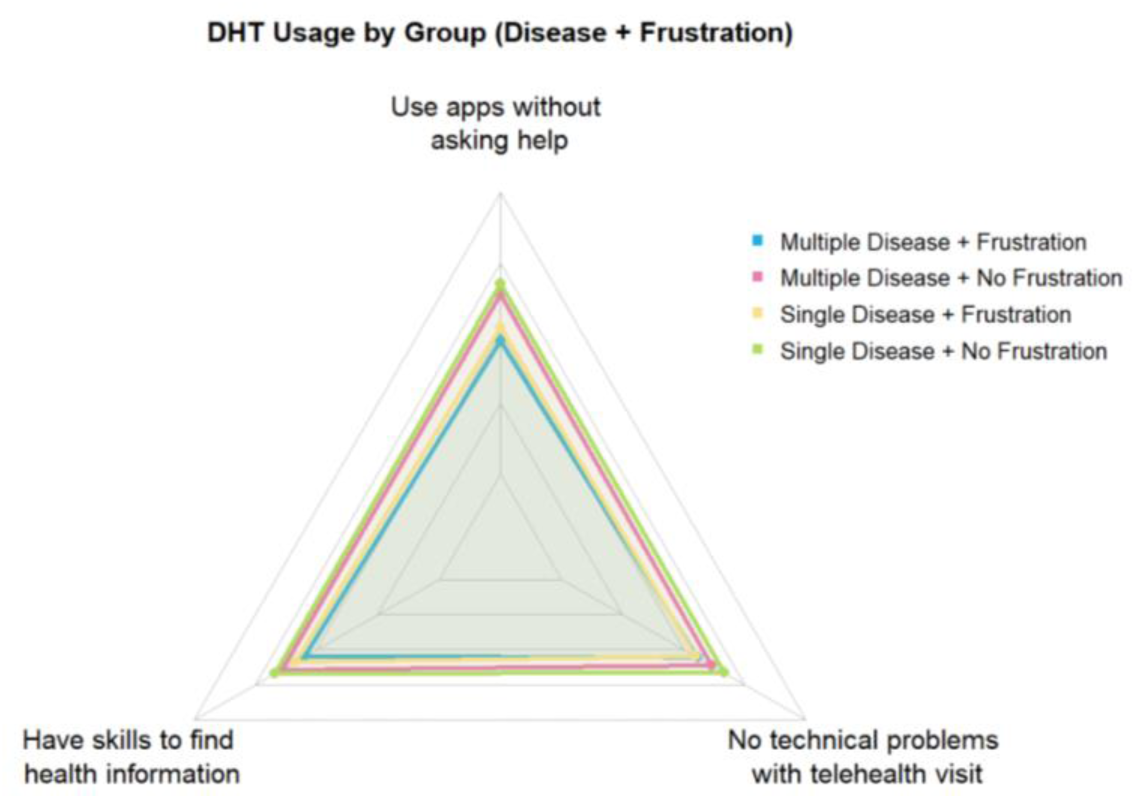
Digital health literacy and operational competence profiles by multimorbidity status and frustration status

**Figure 5b.**
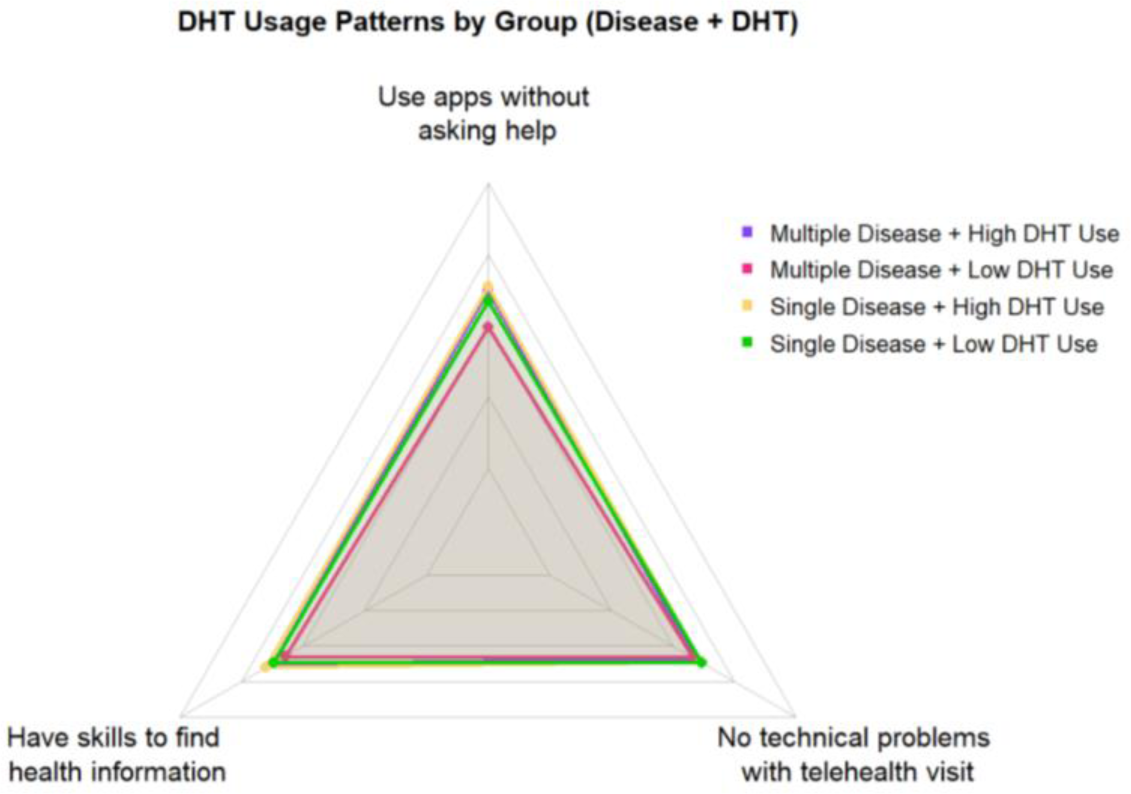
Digital health literacy and operational competence profiles by multimorbidity status and digital health technology usage intensity

Figure 5a reveals clear gradients in self-efficacy and technical competence when participants are grouped by diseases and frustration. The group with a single condition and no frustration exhibited the highest proficiency across all three domains, forming the largest radar area and indicating strong digital health capabilities. They were followed closely by the multimorbidity but not frustrated group, suggesting that the absence of frustration is strongly associated with higher perceived digital capability, even in the presence of multiple chronic conditions. The two frustrated groups (single condition with frustration and multimorbidity with frustration) showed relatively lower scores across all dimensions, with the multimorbidity and frustrated group demonstrating the lowest overall digital literacy profile. This pattern underscores frustration as a key marker of reduced digital competence, compounded by clinical complexity.

Figure 5b revealed a nuanced pattern. For two dimensions—the skills to use apps independently and to find health information online—a consistent gradient was observed: single condition with high DHT use > multimorbidity with high DHT use > single condition with low DHT use > multimorbidity with low DHT use. This gradient suggests that technology adoption level was associated with self-reported digital literacy and that these skills may be enhanced through regular technology use. However, both single condition groups (high and low DHT use) reported similar and higher levels of problem-free telehealth experiences, substantially outperforming both multimorbidity groups, which clustered at lower levels regardless of their DHT adoption intensity.

## DISCUSSION

This study employed a multidimensional analytical approach to examine the complex interrelationships between multimorbidity, DHT use, and user frustration, revealing key patterns that advance our understanding of digital engagement among populations with chronic diseases.

Our analyses demonstrated that individuals with multiple chronic conditions report higher overall levels of frustration with DHTs despite using more types of technologies than those with single conditions. This paradox can be understood through the lens of cumulative burden. The current DHT ecosystem is predominantly characterized by condition-specific applications and platforms, which forces patients with complex care needs to navigate multiple non-integrated devices and applications to manage their health comprehensively.[7] This fragmentation creates substantial coordination demands, increases the cognitive load required to synthesize information across platforms, and amplifies the risk of technical difficulties, duplicate data entry, and conflicting care recommendations. However, our finding that greater intensity of DHT adoption was independently associated with reduced frustration suggests that this relationship is not linear or unidirectional. The observation that individuals with multimorbidity who were highly engaged with DHTs (≥ 6 technologies) exhibited frustration levels comparable to their healthier counterparts with similar technology use indicates that technology proficiency and the development of integrated personal digital health ecosystems may effectively mitigate the potential burden of technology use.[15] This pattern may reflect that individuals who successfully integrate multiple technologies into their routines develop enhanced digital literacy and self-efficacy; or alternatively, it may indicate that those with lower frustration tolerance are more likely to persist in adopting additional technologies. Longitudinal research is needed to disentangle these temporal relationships and identify the mechanisms through which technology proficiency develops.[16]

Another central finding of our study concerns the powerful moderating role of SES in shaping digital health engagement outcomes. Income and education emerged as foundational determinants of both technology adoption patterns and user experience quality.[17] Individuals with higher SES were significantly more likely to adopt multiple DHTs and reported substantially lower frustration levels, irrespective of their multimorbidity. This persistent socioeconomic gradient, which was evident across multiple analytical approaches and remained robust in fully adjusted models, suggests that socioeconomic barriers may represent more fundamental determinants of digital health engagement than clinical factors alone.[18] The mechanisms underlying this socioeconomic gradient are likely multifactorial and include differential access to high-quality devices and reliable internet connectivity, variations in baseline digital literacy and technical skills, differences in the availability of informal support networks for troubleshooting technical problems, and potential disparities in the quality and usability of DHTs provided through different healthcare settings or insurance plans. Consequently, unless DHTs are intentionally designed with equity as a foundational principle, they risk widening pre-existing health disparities by creating additional structural barriers for socioeconomically marginalized populations.[17] This finding has critical implications for policy and implementation: digital health initiatives must incorporate explicit equity metrics, provide robust technical support infrastructure, and ensure that DHT design and deployment strategies actively address rather than perpetuate socioeconomic disparities.

The interaction between multimorbidity and DHT use revealed distinct digital engagement phenotypes with important practical implications for targeted interventions. We identified a particularly informative phenotype comprising individuals with multimorbidity and high DHT engagement (Group 4), who demonstrated frustration levels comparable to their healthier counterparts with similar technology use patterns (Group 2). This pattern suggests that technology proficiency and the successful integration of multiple DHTs into cohesive personal digital health ecosystems may effectively mitigate the digital burden typically associated with complex care needs. These “successful adopters” may benefit from characteristics such as higher baseline digital literacy, stronger self-efficacy, access to better technical support, or exposure to more user-friendly and interoperable systems. Conversely, we identified a high-risk phenotype characterized by older adults with multimorbidity, lower SES, and limited technology adoption (Group 3). This profile demonstrated the highest frustration levels (60.3%) and aligns with documented barriers such as limited digital literacy, physical limitations (e.g., visual impairment, reduced manual dexterity), cognitive challenges (e.g., reduced working memory, slower processing speed), and practical difficulties with fundamental operations such as connecting to the internet, troubleshooting technical problems, or navigating complex interfaces.[19] These individuals are at highest risk for digital exclusion—a phenomenon where marginalized communities with lower digital literacy and internet access are further disadvantaged by technology-dependent healthcare delivery models.[20] This phenotypic stratification highlights how technology adoption and proficiency may empower some patients with complex conditions, while simultaneously underscoring the urgent need for targeted interventions to support vulnerable subgroups as healthcare systems become increasingly digitalized.

The radar chart analyses provided insights into distinct dimensions of digital literacy and operational competence. While overall frustration with DHTs primarily related to general digital self-efficacy and confidence in using asynchronous tools (e.g., health apps and online information seeking), multimorbidity status specifically predicted technical difficulties during synchronous telehealth visits.[21] This dissociation suggests several important conclusions. First, frustration with DHTs appears to stem more from practical operational challenges than from inherent technology complexity per se. These operational challenges include navigating non-intuitive interfaces, managing interoperability issues between disparate systems, coordinating information across multiple platforms, and coping with inadequate training tailored to individual needs and limitations.[22] Second, the finding that multimorbidity predicts telehealth technical difficulties independently of overall DHT adoption suggests that synchronous video-based encounters may impose unique cognitive, physical, or technical demands that are particularly challenging for individuals managing multiple conditions. These demands may include the need to manage medication schedules and symptoms during appointments, coordinate care across multiple specialists simultaneously, communicate complex medical histories under time pressure, or troubleshoot audio-visual problems in real-time while experiencing health-related stress. These findings highlight a critical need for a dual-pronged approach: (1) development of simplified, more intuitive user interfaces that reduce cognitive load and accommodate physical limitations, employing principles of universal design, and (2) implementation of personalized, competency-based training programs that address specific skill gaps and the physical or cognitive limitations common among patients with high multimorbidity burden. Additionally, telehealth platforms may require specialized optimization, such as pre-call technical checks, simplified connection protocols, asynchronous pre-visit information gathering, and dedicated technical support hotlines, to better accommodate patients with complex chronic conditions.[23]

The non-significance of race/ethnicity, sex, and health insurance status in the fully adjusted frustration models, while age, income, and education remained significant, suggests that digital divides may be evolving from primarily access-based disparities toward proficiency-based disparities. As digital technologies become more ubiquitous and internet connectivity improves, the differential ability of individuals to develop and effectively apply digital skills is emerging as the primary driver of digital health inequality.[24] This transition is consistent with scholarship on the “second-level digital divide,” which posits that disparities in how people use technology (skills, self-efficacy, beneficial use patterns) may be more consequential than disparities in whether they have access to technology.[25] However, this interpretation requires several important caveats. First, the HINTS survey is administered primarily through digital platforms (mail and web-based), which may systematically underrepresent populations with severely limited digital access or literacy, potentially masking the true extent of access-based disparities. Second, our finding of persistent SES gradients (income and education) suggests that structural inequities continue to shape digital health engagement, likely through mechanisms that encompass both access and proficiency dimensions.[26] Third, the operational challenges highlighted by our findings, such as difficulties navigating complex interfaces and managing multiple non-integrated systems, align with the concept of digital exclusion among older adults and other vulnerable groups who often face compounded barriers related to meaningful digital participation.[27]

These findings underscore that while improving digital literacy and skills training is essential, these efforts must be complemented by continued attention to structural barriers (affordability, connectivity infrastructure, device quality) and by fundamental improvements in DHT design that reduce the baseline skill requirements for effective use. The goal should not be merely to train more patients to use complex systems, but to create simpler, more intuitive systems that accommodate diverse user needs and capabilities from the outset. These findings also highlight the need for multi-level interventions. At the system level, healthcare organizations and policymakers should prioritize the development of interoperable platforms that reduce the burden of managing multiple applications,[28] establish universal design principles that accommodate diverse user capabilities, and implement equity metrics to monitor and address digital health disparities.[29] At the individual level, targeted interventions should focus on personalized digital literacy training, peer support programs, and accessible technical assistance—particularly for older adults with multimorbidity and low SES who are at highest risk for digital exclusion. Intervention studies are needed to test strategies for reducing frustration and improving digital health equity.[30] Promising approaches include personalized digital literacy training programs tailored to individual skill levels and learning styles, peer support and mentorship programs connecting successful adopters with those struggling with technology, technical support hotlines with trained specialists who understand the unique needs of older adults and those with chronic conditions, and co-design processes that involve patients with multimorbidity in the development and iterative refinement of DHTs to ensure they meet real-world needs.[31]

### Limitations

This study has several limitations. First, the cross-sectional design cannot provide causal inference regarding the observed relationships. Self-reported technology use may be subject to recall bias and social desirability bias. Second, the DHT threshold (i.e., 6 technologies), while statistically derived, its clinical relevance requires practical validation in diverse populations. Third, as the HINTS survey is administered primarily through digital platforms, it may systematically underrepresent populations with limited digital access or literacy, potentially underestimating the true digital divide and the frustration burden among the most vulnerable groups. Since the survey was conducted exclusively in the United States, the findings’ generalizability may be limited by the cultural and healthcare system context, requiring validation in other national settings. Future research could employ longitudinal designs to track how technology adoption patterns evolve with changing health status, and incorporate objective measures of digital engagement, such as platform usage metrics and device-collected data, to complement self-reported measures.

## CONCLUSION

This study provides empirical evidence that digital health engagement is not merely a function of clinical need or technology availability, but rather reflects complex interplays between socioeconomic resources, technology proficiency, and clinical complexity. The identified phenotypes may offer a novel framework for identifying vulnerable subgroups that would benefit most from tailored digital health strategies and for understanding the mechanisms through which technology may empower some patients while excluding others. The goal should be to create digital health ecosystems that genuinely reduce rather than merely redistribute the burden of chronic disease management. This requires moving beyond a narrow focus on technology adoption rates to embrace a more comprehensive vision of digital health equity that attends to user experience, operational competence, and the structural determinants of meaningful engagement. Only through such patient-centered approaches can digital health technologies fulfill their potential to support improved outcomes and reduced burden for all patients with multimorbidity.

## CONFLICTS OF INTEREST

None.

## CONTRIBUTION STATEMENT

JY designed the study, contributed to the data analyses, and the writing of the manuscript. HC contributed to data analyses and the writing of the manuscript. All authors read and approved the final version of the manuscript.

## DATA AVAILABILITY

Data from the Health Information National Trends Survey (HINTS 7) are publicly available from the National Cancer Institute at https://hints.cancer.gov/

## FUNDING

None.

## Notes

### Competing Interest Statement

The authors have declared no competing interest.

### Author Declarations

National Cancer Institute at https://hints.cancer.gov/

### Summary of Updates

The author's affiliation was updated.

